# Serious mental health diagnoses in children on the Child Protection Register: a record linkage study

**DOI:** 10.1101/2023.10.13.23296488

**Authors:** William P Ball, Caroline Anderson, Corri Black, Sharon Gordon, Michael Lackenby, Martin Murchie, Bārbala Ostrovska, Katherine O’Sullivan, Helen Rowlands, Magdalena Rzewuska Díaz, Jessica E Butler

## Abstract

**Purpose:** Children with experience of maltreatment, abuse or neglect are known to have a higher prevalence of poor mental health. Child Protection Services identify children most at risk of harm and in need of intervention. Mental healthcare usage in this population is not well understood as registration data is not routinely linked to health records.

**Methods:** We undertook data linkage to describe the population on the register, their mental healthcare usage and to calculate age- and sex-specific incidence rates of mental health outcomes. We analysed records from the Aberdeen City Council Child Protection Register and for mental health prescribing and referrals to child and adolescent mental health services (CAMHS) for the NHS Grampian region between 1^st^ January 2012 and 31^st^ December 2022.

**Results:** We identified 1,498 individuals with a Child Protection Register registration, of which 70% were successfully matched to health records. 20% of registrations occurred before birth and the median age of registration was 3 years. 10.1% of children with a registration ever received a mental health prescription, 5.1% for treatment of attention deficit hyperactivity disorder and 1.7% for treatment of depression. 18.9% received a referral to specialist outpatient Child and Adolescent Mental Health Services. Age- and sex- standardised incidence rates for mental health prescribing and referrals are higher for children with a child protection registration compared to the general population.

**Conclusion:** Children identified as being at significant risk of harm and involved with child protection services are at greater risk of seeking or receiving professional mental health support than their peers. Clinical services should investigate additional ways to support this population’s mental well-being as a priority. Efforts to reduce the exposure of children to potentially harmful environments at a societal level should also be pursued.

## Background

Childhood experiences of maltreatment, abuse or neglect can have serious harmful effects on physical and mental health [1–4] and present major and longstanding population health issues worldwide [5]. These experiences are known to be strongly associated with poorer physical and mental health outcomes [6, 7] which can have effects across the life course [8, 9]. In response to these challenges, governments around the world have implemented statutory frameworks and Child Protection Systems to fulfil their duty to reduce the risk of harm [10].

In Scotland and the rest of the United Kingdom, a Child Protection Register (CPR) is a confidential list of children in a local authority area who have been identified as being at risk of significant harm and in need of intervention. In addition to appearing on the list, a Child Protection Plan (CPP) will also be in place which outlines how checks on child welfare will be carried out, what measures are required to reduce risk and what support is to be offered to the family [11].

The most common reasons cited for children entering the register include concerns about the risk of harm due to domestic abuse, neglect, parental mental health problems or parental substance abuse [12]. There are between 3,000 and 4,500 child protection registrations across Scotland each year, with the vast majority for children who have not had a previous registration [13].

This group of children are likely to be at increased risk of suffering from poor mental health either in childhood or later life, although this is currently under-researched. However, epidemiological studies exploring mental health in these populations are relatively rare, with one meta-analysis also reporting heterogeneous findings in the observed prevalence of mental disorders [14]. Other research found that almost half of children with experience of child welfare services had clinically significant emotional or behavioural problems, as well as an increased likelihood of receiving mental health care [15]. Another study found an increased risk of receiving a psychotropic prescription, hospital admission or emergency presentation for self-harm compared to children without interaction with social services [16]. Conducting this kind of analysis is important to inform the design and provision of mental healthcare services, as well as social policy aimed at reducing the prevalence of poor mental health.

This type of research is complicated by individual-level information on child protection proceedings and electronic health records being held by Local Authorities and Health Boards respectively, without a common unique identifier. This study links information about children on the Aberdeen City Council Child Protection Register to their electronic health records, specifically for specialist outpatient child and adolescent mental health services (CAMHS) referrals and community mental health prescribing. We have the benefit of access to records for the total population living in the NHS Grampian health board (for health records) and in Aberdeen City Council (for CPR registrations) during the study period, which allows for direct comparison between children with registration and those without.

We present a detailed epidemiological description of children on the CPR and their mental health care use.

## Methods

### Study Design

This is a retrospective dynamic cohort study examining electronic health records linked to administrative data from child protection services.

### Setting

This study setting is the NHS Grampian Health Board region in the North East of Scotland which covers 3 Local Authority areas (Aberdeen City, Aberdeenshire and Moray Councils). Child Protection Register data relates only to children living within the Aberdeen City Council boundary area. The study period is between 1^st^ January 2015 and 31^st^ December 2022. In each year of the study period, there were roughly 110,000 individuals living in NHS Grampian aged under 18 years [17]. Accounting for the study period window (i.e. individuals leaving and newly entering the cohort), 145,000 individuals were eligible for inclusion based on mid-year population estimates.

### Data Sources

The Aberdeen City Council Child Protection Register (ACCCPR) records a list of all children living in the area who are formally recognised as being at risk of significant harm. Child and Adolescent Mental Health Services (CAMHS) referrals data is an NHS Grampian dataset used to report official waiting list statistics nationally. The Prescribing Information System (PIS) provides records for all medicines prescribed or dispensed in the community setting. The Scottish Index of Multiple Deprivation (SIMD) 2020 version 2 provides an area measure of deprivation based on place of residence. Rates have been calculated for outcomes using official mid-year population estimates for the NHS Grampian and Aberdeen City Council regions [18].

Data linkage and management were conducted by Grampian Data Safe Haven (DaSH) [19] staff on a secure server using accredited procedures to collect, link and pseudonymise patient-level records. Access to de-identified records was allowed through the Trusted Research Environment (TRE) for only named and approved researchers and outputs in the form of aggregate summary data, tables or figures were subject to disclosure control procedures to reduce the risk of identifiable information being shared.

#### Community Health Index Matching

The common unique identifier in all health datasets in this study is the Community Health Index (CHI) number, held in the CHI register [20], which is a list of all patients in NHS Scotland. The ACCCPR is maintained by Aberdeen City Council (not the NHS) and does not have a CHI number attached to any records. To facilitate individual-level data linkage, CHI numbers were matched from NHS Grampian CHI Registers (from 2012 and 2021) to records in the ACCCPR using deterministic record linkage based on matching combinations of forename, surname, date of birth and postcode of residence between the two data sources.

The matching takes place in a sequential process, attempting to match on different criteria. There were 12 match types or ’blocks’ in total, with 4 considered ‘high quality’, and 8 of ‘lower quality’ (table 1). Those with single incidence ’high quality’ matches in the first 4 blocks were excluded from further matching attempts. The matches were scored, and a score threshold was determined for inclusion from the lower-quality matches. These methods were based on an approach published elsewhere [21].

**Table 1.** CHI Matching Criteria for automated stepwise matching. Note: Unless otherwise indicated (e.g. blocks 9 and 10) all searches were against the NHS Grampian CHI register from 2021

| Quality | Block | Family Name |  | Forename | Date of Birth | Postcode (Current OR Previous) |
| --- | --- | --- | --- | --- | --- | --- |
|  |  | Current | Previous |  |  |  |
| High | 1 | X |  | X | X | X |
| High | 2 |  | X | X | X | X |
| High | 3 | X |  | X | X |  |
| High | 4 |  | X | X | X |  |
| Lower | 5 |  |  | X | X |  |
| Lower | 6 | X |  |  | X |  |
| Lower | 7 | X (Current OR Previous) |  |  | X |  |
| Lower | 8 |  |  |  | X | X |
| Lower | 9 |  |  |  | X | X (postcode from NHS Grampian patient history) |
| Lower | 10 |  |  |  | X | X (postcode from 2012 CHI register) |
| Lower | 11 |  |  |  | X |  |
| Lower | 12 | Split Names | Split Names | Split Names |  |  |

### Participants

#### Inclusion Criteria

All people living in NHS Grampian Health Board during the study period who were aged 17 years or younger between 1^st^ January 2012 and 31^st^ December 2022 and:

− Were registered on the Aberdeen City Council Child Protection Register, **OR**
− Received a prescription for medications to treat mental health conditions, **OR**
− Were referred to/attended specialist outpatient CAMHS.

The upper age limit has been chosen to align with the population who can ordinarily be referred to specialist outpatient CAMHS (under 18). There is no lower age limit because children can appear on the child protection register pre-birth.

### Variables

Presence on the CPR has been identified by dates of registration or deregistration from the ACCCPR. For calculation of prevalence proportions and incidence rates, the group listed on the Child Protection Register includes children whose outcome occurs before, during or after their registration – i.e. they are children who have ever had a registration. Referrals to specialist outpatient CAMHS are defined as those made to Scottish ‘tier 3’ services, i.e. those provided by locality teams based in a hospital. The date of referral was used to identify that a referral was made during the study period.

Mental health-related prescriptions have been identified by the British National Formulary (BNF) item code recorded in PIS or by item name where no item code is available. Medications in the following BNF sections have been included: 4.1) Hypnotics and Anxiolytics, 4.2) Drugs used in psychoses and related disorders, 4.3) Antidepressant drugs, 4.4) Central nervous system stimulants and drugs used for ADHD, and 4.10) Drugs used in substance dependence [22]. These classes of medication align with the classification of mental health medications applied elsewhere [23, 24]. A full list of included medications can be found in supplementary table S3.

A count of children without a CPR registration, mental health prescription or specialist outpatient referral has been derived from annual mid-year population estimates by year of age and sex [18].

### Statistical Methods

We describe the social and demographic characteristics of the entire population on the register. This includes descriptive summaries of age, sex and length of time on the register over the entire study period and by year. We describe mental health prescribing and outpatient referrals to CAMHS [24] for the population who have ever appeared on the Child Protection Register. We have calculated the prevalence of mental health prescriptions (by class) and specialist outpatient referrals over the entire study period and by year.

We also compare the mental health care use for all children in NHS Grampian with the population listed on the Child Protection Register. We calculate age- and sex-specific incidence rates of mental health prescription (overall and by medication class) and referral to CAMHS for children on the child protection register and those with no registration [25]. Year of age has been grouped into four categories (0-4 years, 5-9, 10-14 and 15-17). Denominators for incidence rate are per 1,000 person-years and annual mid-year population statistics [18] have been used to calculate the population at-risk for those without a CPR registration.

CPR status was determined by a binary indicator for individuals who have ever had a registration. Denominators for this group were determined by the number of individuals with a CPR registration who were at risk of an outcome during a given year, by age group and sex. Incidence rates are expressed per 1,000 person-years and can be interpreted as analogous to the number of outcomes you might observe after following 1,000 individuals in this category for a single year.

We calculate Incidence Rate Ratios (IRRs) and 95% Confidence Intervals (CIs) to compare age- and sex-specific incidence rates for mental health outcomes between children with and without a CPR registration [26]. The IRR is expressed as the relative difference between the incidence rate per 1,000 person-years in the CPR group and the non-CPR group. An IRR less than 1 indicates that the incidence rate in the CPR group is lower. An IRR equal to 1 indicates no difference in incidence rates between CPR groups. An IRR greater than 1 indicates that the rate in the CPR group is higher than in the non-CPR group.

## Results

### Cohort Description and CHI matching – The Aberdeen City Council Child Protection Register

Table 2 provides a summary of the Aberdeen City Council Child Protection Register between 1^st^ January 2012 and 31^st^ December 2022. During this period 1,498 individual children received 1,719 total registrations. The annual mean for new registrations is 171 and for individuals is 170. Table 3 describes the children who could and could not be matched to their health records. Overall, 1,029 (69%) individuals on the register were successfully CHI matched and this accounts for 70% (n = 1,202) of all registration records.

**Table 2.**
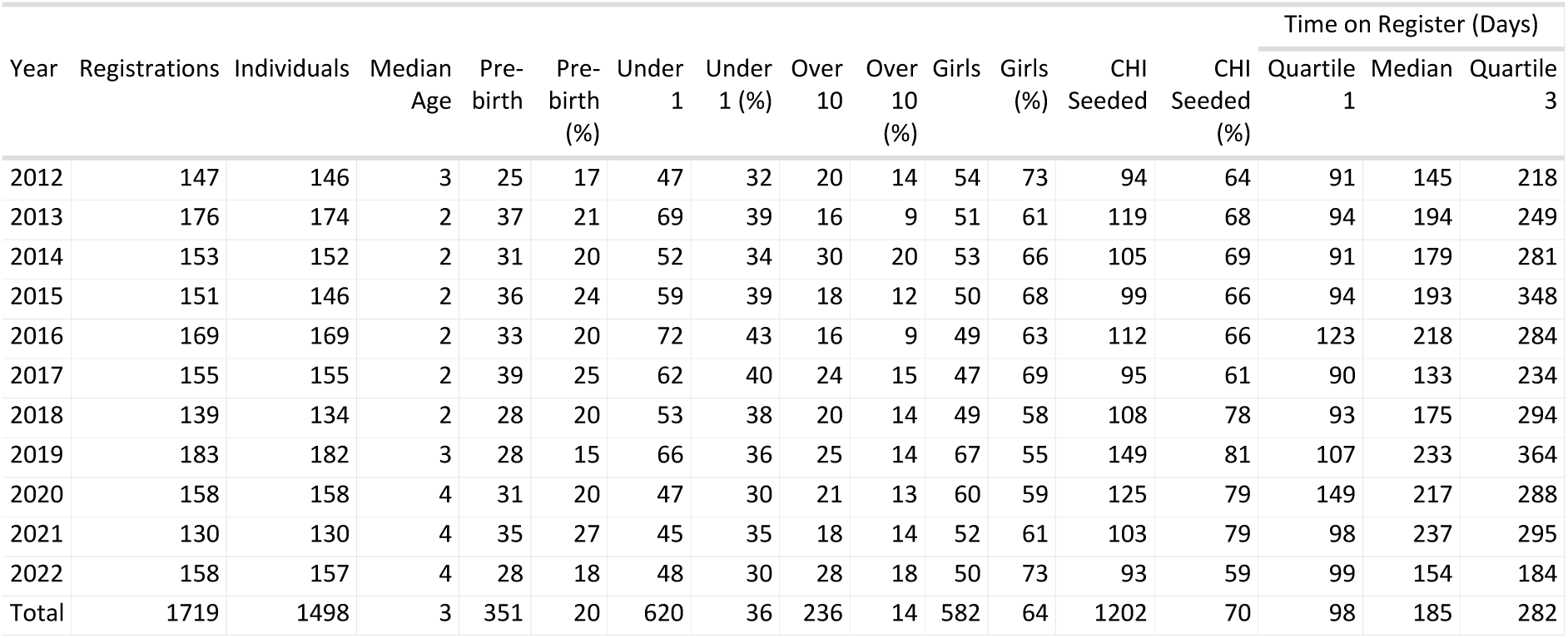
Summary of age, sex, CHI matching status and length of time listed on the Aberdeen City Council Child Protection Register, annually and in total (2012 - 2022). Note: Sex was only available for successfully CHI-matched records – proportions of girls reported here relate only to those with a CHI.

**Table 3.** Descriptive summary of individuals on the ACCCPR who have been successfully CHI-matched and unmatched. Note: for the first registration.

| CHI Matched | Individuals | Mean Age (Years) | Median Age (Years) | Pre-birth | Pre-birth (%) | Under 1 | Under 1 (%) | Over 10 | Over 10 (%) | Time on Register (Days) |  |  |
| --- | --- | --- | --- | --- | --- | --- | --- | --- | --- | --- | --- | --- |
|  |  |  |  |  |  |  |  |  |  | Quartile 1 | Median | Quartile 3 |
| 0 | 469 | 3.6 | 0 | 161 | 34 | 252 | 54 | 42 | 9 | 91 | 150 | 254 |
| 1 | 1029 | 4.4 | 3 | 190 | 18 | 356 | 35 | 157 | 15 | 99 | 191 | 286 |

20% of all registrations (n = 351) were made for children before their date of birth, but this proportion was almost double in records which were not matched to a CHI (34% vs. 18% in matched). For registrations following birth, the median age at registration was 3 years, ranging annually between 2 (Quartile 1) and 4 (Quartile 3) years of age. For records without a matched CHI the median age was 0 years (i.e. after birth, but aged less than 1 year).

The median number of days on the register for each registration was 185 (Q1: 98, Q3: 282). The smallest annual median time on the register was 133 days for registrations starting in 2017 and the largest was 237 days for registrations starting in 2021.

### Mental Health Prescribing and Specialist Outpatient Referrals

Of individuals who have appeared on the ACCCPR between 2012 and 2022, 10.1% (n = 161) received at least one mental health prescription during the study period. 5.1% (n = 76) were prescribed a medicine used to treat ADHD and 1.7% (n = 25) were prescribed a medicine to treat depression (Table 4). CAMHS referrals were made for 18.9% (n = 283) of individuals with a CPR registration. Annual counts and proportions are presented by year of first CPR registration and are generally higher for those registered earlier in the study period.

**Table 4.** Prevalence over time of Prescribing (For all Mental Health medications and specifically for the treatment of ADHD and Depression) and Specialist Outpatient Referrals, by year of 1st registration.

| Year | Individuals | Prescribing |  |  |  |  |  | CAMHS Referrals |  |
| --- | --- | --- | --- | --- | --- | --- | --- | --- | --- |
|  |  | With Prescription | Prescription (%) | ADHD Prescription | ADHD (%) | Antidep Prescription | Antidep (%) | With Referral | Referral (%) |
| 2012 | 146 | 22 | 15.1 | 10 | 6.8 | 6 | 4.1 | 40 | 27.4 |
| 2013 | 174 | 23 | 13.2 | 18 | 10.3 | <5 | - | 38 | 21.8 |
| 2014 | 152 | 16 | 10.5 | 6 | 3.9 | 5 | 3.3 | 33 | 21.7 |
| 2015 | 146 | 15 | 10.3 | 8 | 5.5 | <5 | - | 22 | 15.1 |
| 2016 | 169 | 21 | 12.4 | 13 | 7.7 | 5 | 3 | 28 | 16.6 |
| 2017 | 155 | 7 | 4.5 | <5 | - | <5 | - | 20 | 12.9 |
| 2018 | 134 | 10 | 7.5 | 6 | 4.5 | <5 | - | 19 | 14.2 |
| 2019 | 182 | 14 | 7.7 | 8 | 4.4 | <5 | - | 22 | 12.1 |
| 2020 | 158 | 6 | 3.8 | <5 | - | <5 | - | 16 | 10.1 |
| 2021 | 130 | 8 | 6.2 | <5 | - | <5 | - | 13 | 10 |
| 2022 | 157 | 10 | 6.4 | <5 | - | <5 | - | 16 | 10.2 |
| Total | 1498 | 152 | 10.1 | 76 | 5.1 | 25 | 1.7 | 283 | 18.9 |

### Age and Sex-Specific Incidence Rates

Table 5 shows incidence rates per 1,000 person-years for mental health outcomes by sex, age group and by child protection registration or not (CPR Status). CPR status is determined by whether an individual has ever had a registration regardless of whether their outcome was before or after registration.

**Table 5.** Outcome incidence rates (per 1,000 person-years) grouped by age, sex and CPR status.

| Female |  |  | MH Prescriptions | ADHD Prescriptions | Antidep Prescriptions | CAMHS Referrals |
| --- | --- | --- | --- | --- | --- | --- |
| Age | 0-4 | NHS Grampian | 1.5 | - | - | 2.3 |
|  |  | CPR | - | - | - | 12.3 |
|  | 5-9 | NHS Grampian | 5.8 | 1.9 | 0.3 | 10.8 |
|  |  | CPR | 16.5 | 8.7 | - | 19.4 |
|  | 10-14 | NHS Grampian | 11.7 | 1.6 | 4.7 | 25.9 |
|  |  | CPR | 18.7 | - | - | 43.7 |
|  | 15-17 | NHS Grampian | 45.4 | 1.7 | 29.6 | 35.4 |
|  |  | CPR | 67.3 | - | 39.7 | 63.7 |
| Male |  |  | MH Prescriptions | ADHD Prescriptions | Antidep Prescriptions | CAMHS Referrals |
| Age | 0-4 | NHS Grampian | 3.0 | - | 0.1 | 5.3 |
|  |  | CPR | 10.0 | - | - | 21.4 |
|  | 5-9 | NHS Grampian | 18.6 | 8.9 | 0.5 | 17.1 |
|  |  | CPR | 50.4 | 24.9 | - | 35.8 |
|  | 10-14 | NHS Grampian | 15.2 | 4.9 | 2.7 | 17.7 |
|  |  | CPR | 39.0 | 19.2 | - | 37.7 |
|  | 15-17 | NHS Grampian | 24.1 | 1.9 | 12.4 | 19.4 |
|  |  | CPR | 28.0 | - | 38.8 | 25.6 |

Incidence rates for any mental health prescription increase with age and are higher in males than females, apart from in the oldest age group. For both males and females and at each age group, incidence rates for any prescription are higher in the CPR group.

Where it has been possible to calculate incidence rates for ADHD prescriptions (i.e. where the number of outcomes is not less than 5 for disclosure purposes), they are highest in males aged 5-9 (IR: 24.9 per 1,000 person-years). Rates of ADHD prescription are higher in males than females in each age group. Females aged 5-9 with a CPR registration have almost the same rate of ADHD prescription as males of the same age without a registration (CPR: 8.7 per 1,000 person-years, Non-CPR: 8.9).

Antidepressant prescription rates are higher in females than males and highest in the oldest age group. Males and females in the 15-17 year age group with a CPR registration have the highest rates of antidepressant prescribing (Males: 38.8 per 1,000 person-years, Females: 39.8) and the male CPR rate is notably higher than the female non-CPR rate (26.6 per 1,000 person-years).

CAMHS referral incidence rates are higher for children with a CPR registration in both sexes and at each age group (Table 5 and Figure 1). Rates for females increase with age and are highest in the oldest age group (Non-CPR: 35.4 per 1,000 person-years, CPR: 63.7). Rates for males are higher than for females in younger ages but have a smaller increase by age. The highest incidence rates for males on the CPR are in the 10-14 years age group (37.7 per 1,000 person-years).

**Figure 1.**
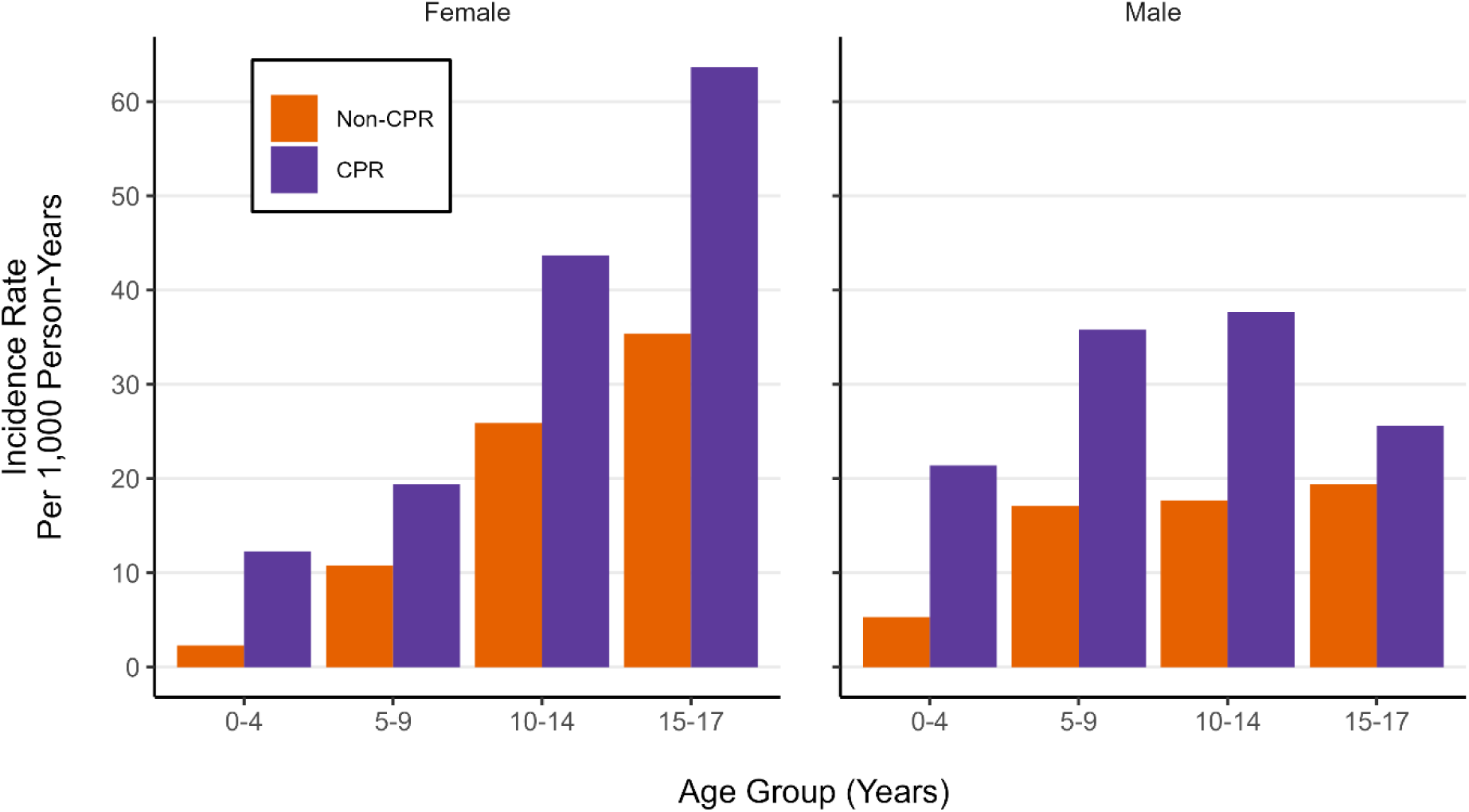
Incidence Rates for CAMHS Referrals per 1,000 Person-years.

### Incidence Rate Ratios

Figure 2 shows the incidence rate ratios between children with and without a CPR registration for mental health outcomes, by sex and age group. It was only possible to calculate the ratio where there were incidence rates for both CPR and non-CPR groups. Children with a CPR registration have higher incidence rates in each outcome, in both sexes and in each age group, although 95% Confidence Intervals overlap 1 in some categories. The largest relative difference between incidence rates is for CAMHS referrals in females aged 0-4 years, where children with a CPR registration have a rate that is 5.4 times higher (95% CIs: 2.9 – 9.1). The largest absolute difference is for mental health prescriptions in males aged 5-9 years, where children with a CPR registration have an additional 31.8 prescriptions per 1,000 person-years. With a rate of 50.4 prescriptions per 1,000 person-years, these children have a 2.7 times higher incidence rate (95% CIs: 2.1 – 3.4).

**Figure 2.**
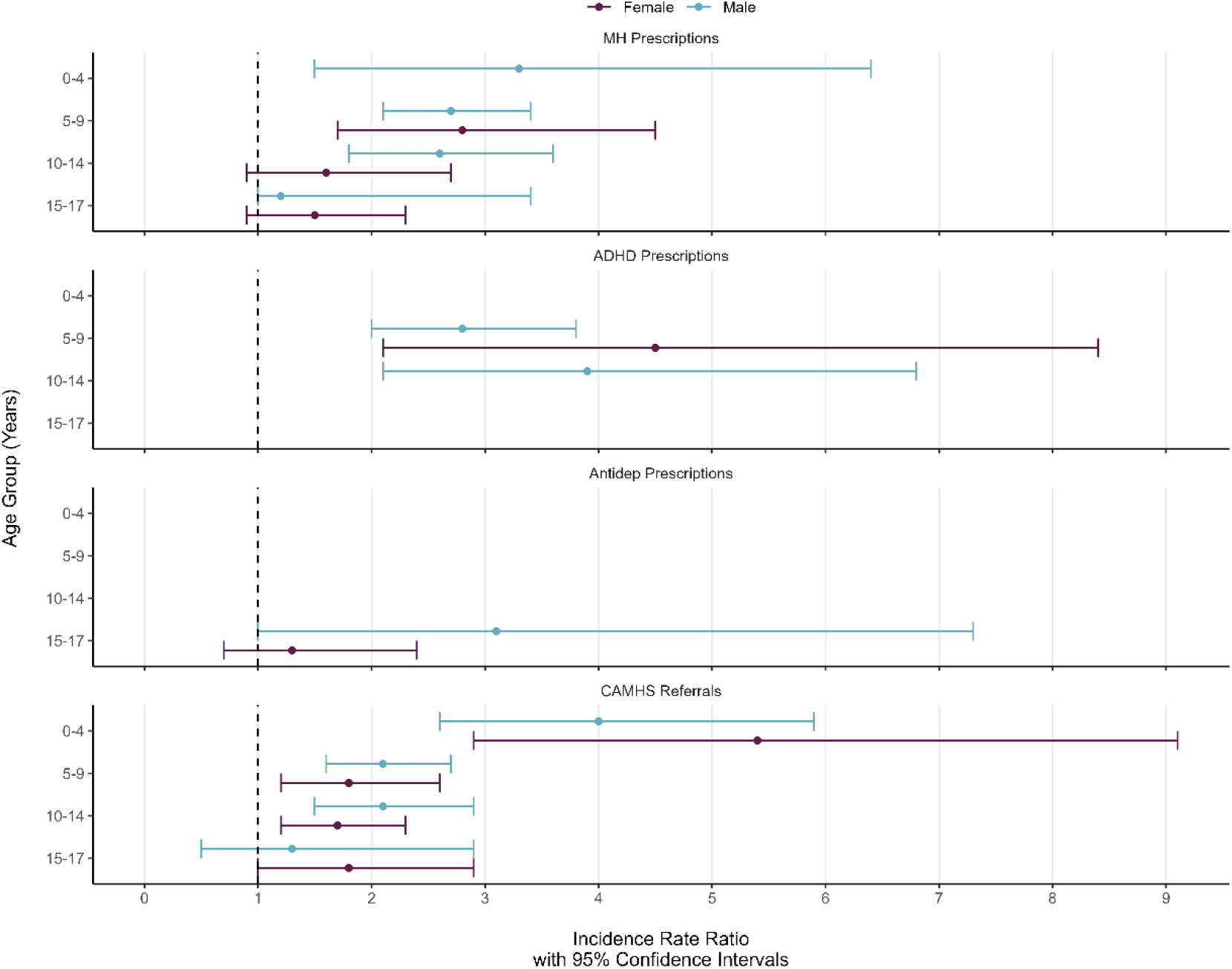
Incidence Rate (per 1,000 person-years) Ratio, and 95% Confidence Intervals, for children with and without a CPR registration, by sex and age group. Note: The dashed line indicates where there is no difference between incidence rates.

## Discussion

### Principle Findings

#### Mental Health Outcomes

We found that children who have ever had a CPR registration were at a greater risk of receiving a mental health prescription (of any kind), specifically for the treatment of ADHD or depression, as well as referrals to specialist outpatient CAMHS. The increased risk was observed in all age groups from 0-17 years and for both males and females. The increased risk, in relative terms, was greatest in the youngest age group.

We found that almost 1 in every 5 (18.8%) children with a CPR registration during the study period had also received a referral to specialist outpatient CAMHS. 1 in 10 (10.1%) had received a mental health prescription, with 1 in 20 (5.1%) for treatment of ADHD and almost 1 in 60 (1.7%) for depression. However, due to the young age at which most children receive a CPR registration, these outcomes are patterned by the time at which they entered the cohort. Proportions of outcomes are higher in children with a registration in the early years of the study period as they have had more time to experience an outcome. Age is also an important consideration as treatments for ADHD are more likely to be described in younger ages and antidepressants in older teenagers. Given the very young age at which people receive a CPR registration and patterns of prescribing by age, this makes the calculation of age-specific incidence rates essential to comparing risk between the CPR and non-CPR populations.

#### Aberdeen City Council Child Protection Register

We found that a substantial proportion of CPR registrations are for very young children, including many registered before their birth. Trends in annual registrations as well as distributions of age, sex and number of days on the register have remained relatively consistent throughout the study period.

#### CHI Matching

We found that almost one-third of children listed on the ACCCPR could not be matched to their health records and that there appear to be some substantive differences between those who could and could not be matched, at least based on the limited information available to assess this. The children who could not be matched to a CHI record were on average much younger (median 0 years vs. 3 years for those matched) and a higher proportion of unmatched children were registered before their birth. The presence of pre-birth registrations may also explain why the proportion of CHI-matching is lowest in 2022, as these children may not have been allocated a CHI number at the point of data extraction.

### Strengths and Limitations

#### Strengths

This study benefits from total population coverage of health and CHI records for the NHS Grampian region and in the Aberdeen City Council area for the ACCCPR. CAMHS referral data are recorded as part of a statutory requirement for reporting national statistics on patient waiting list times. PIS is the definitive source of information relating to prescriptions dispensed or prescribed in the community. Individual-level linkage of these data sources, including by matching CHI identifiers to the ACCCPR, adds value to these high-quality administrative data sources. This work reveals trends which are not visible in each dataset alone and which are not available in publicly published aggregate statistics. To our knowledge, this is the first linkage of its kind in Scotland, providing previously unknown information but also highlighting challenges that may be addressed in future linkage efforts. Finally, this research has been informed at all stages of the research and publication process by extensive Public and Patient Inclusion and Engagement activities, including topic selection, methods development and interpretation of results.

#### Limitations

The main limitation of this descriptive study is the lack of a common identifier in the data sources which required a deterministic data linkage approach. Although a majority of matched records were deemed ‘high quality’ (i.e. based on matches in forename, surname and date of birth with or without address), a proportion was not and almost one-third of individuals with a registration could not be matched to their health records. Given observed differences between matched and unmatched individuals it is likely that the probability of linkage varies between some population subgroups, which may lead to selection bias influencing observed rates of outcomes. If unmatched children with a CPR registration later have an outcome in the data, it may be recorded in the non-CPR counts/rates. This will lead to an underestimation of the difference between the groups. Another source of potential underestimation is the non-identification (in this data) of children with a CPR registration from outside of Aberdeen City, including residents in the rest of the NHS Grampian Health Board.

Prescribing and specialist outpatient CAMHS referral data are both administrative sources and do not necessarily reflect clinical need/severity (in the case of referral) or the clinical indication (for prescription). Although the British National Formulary (BNF) groups together medications which are intended to be used for similar purposes, some are applicable to treat multiple conditions or can be prescribed ‘off license’ (intended to treat another condition).

### How does it compare with other work?

Variability in the organisation of child protective systems around the world makes direct comparison to the findings of this work somewhat difficult. However, there is an established literature exploring the mental health outcomes of children who have some form of contact with state-run social care/welfare services. This ranges from instances where children have been referred for suspected maltreatment, to being placed in foster or residential care away from their biological parents.

Despite this variability in the populations being studied, there is a concordance among the international literature to find that these children have a higher prevalence of mental health disorders than the general population [1, 14, 27, 28]. Greater involvement (i.e. a higher level of intervention severity) with child protective services has been associated with a higher prevalence of mental health disorders [29], although all levels of involvement had a higher prevalence than the general population. The general pattern aligns well with our finding of a higher risk of receiving a mental health prescription or specialist outpatient referral for children with a child protection registration in this study. A Norwegian study exploring the prevalence of mental health disorders and ADHD in children in child welfare found that both were higher than the general population [30], which appears to complement our findings related to outpatient referrals and classes of medication.

Other studies have also looked at outcomes related to health service use or prescribing. Prescribing of psychotropic medications in youth in child welfare/protective services is higher overall for this population of children, although there is variation based on the frequency of medication use [31] and by state in the United States of America [32]. Although not tested in this study, other research has found higher rates of mental health-related hospitalisation [33] and emergency department attendance [34]. A Scottish study of ‘Looked After Children’ found increased rates of ADHD, depression and self-harm [35]. In Scotland, a child is ‘Looked After’ once they are legally under the care of their Local Authority. This generally includes being placed into foster or residential care but may also include staying at home (with regular social work contact). Looked After Children are taken into care for many of the same reasons that children appear on the Child Protection Register, but the populations will differ as not all children on the CPR will become Looked After. Children may also be removed from the Child Protection Register once they become Looked After and enter care, as their risk of harm may then be sufficiently reduced. Comparing results from this study and a previous study of prescribing and referrals for all children in NHS Grampian [24] shows that many of the trends and patterns are replicated in children on the child protection register, but also that they have increased rates of these outcomes.

### Unanswered Questions and Future Research

Future research should calculate the incidence and prevalence of poor mental health in this population using self-reported measures such as the Strengths and Difficulties Questionnaire (SDQ) [36] or the Warwick-Edinburgh Mental Well-being Scale (WEMWBS) [37] to compare with patterns in mental health prescribing and specialist outpatient referrals. As indicators of service use, the outcomes of interest in this work may not reflect the true need for mental health support. Additional individual-level epidemiological studies, supported by data linkage, can help assess whether patterns of higher risk of mental health service use are due to increased need or increased recognition of need in this population. This work can inform policy aimed at reducing exposure to harm in childhood and around service prioritisation for this population.

This research looked at 10 years of CPR registrations and associated health records. Due to the young age at which children are commonly added to the CPR and the length of our study period, the youngest in the cohort have less opportunity to have an outcome of interest. Future work should explore longer-term outcomes for children listed on the CPR.

This work was able to link a single CPR from Aberdeen City Council to health records. Future work should expand the geographical scope of this project, ideally to achieve national coverage, or at least to include multiple (preferably neighbouring) local authorities. This is particularly pertinent to future research exploring longitudinal or life course outcomes in this population as moving between local authority areas whilst appearing on a CPR is a trigger for de-registration. In addition, children moving into the area may have had a preceding CPR registration elsewhere that is not recognised in the local data. To aid future research in this area, the common identifier for health records should be included in the Child Protection Register and other social work data as standard to enable exact matching approaches to data linkage. This will avoid the requirement for probabilistic record linkage approaches likely to result in incomplete linkages, leading to the underestimation of outcome incidence rates. This may also introduce selection bias into any causal analysis as the likelihood of linkage in population sub-groups may vary [38].

## Conclusions

We demonstrate that children appearing on the child protection register are at increased risk of mental healthcare usage, at each age group, for both sexes and in each of the outcomes measured. We found that age- and sex-based patterns of mental healthcare usage in children with a child protection registration are like their peers without a registration. Future research should prioritise work which can inform policy intervention or service prioritisation to support this population.

This work demonstrates the feasibility of the linkage between administrative Child Protective Services data and health records for research. This work has also highlighted limitations in a probabilistic matching approach. To avoid introducing selection or other sources of bias into future research, a common unique identifier should be included in both data sources and national coverage should be prioritised.

## Supporting information

Supplementary Table 1. CRediT authorship statement

Supplementary Table 2. GRIPP2 PPIE Reporting

Supplementary Table 3. List of medicines

## Contributors

Contributor Roles Taxonomy (CRediT) [39] has been used to make author contributions explicit and is available in the supplementary materials (Table S1) accompanying this publication.

## Ethical Declaration

This project was approved by the North Node Privacy Advisory Committee (NNPAC) (project ID: 6/105/22). NNPAC provides researchers with streamlined access to NHS Grampian data for research purposes and committee approval incorporates approvals from; the project sponsor, institutional ethics committee, the local Caldicott Guardian, and NHS Research & Development. It was not practicable to obtain informed consent from individual patients for this research which uses electronic patient records. Use of this pseudonymised unconsented patient information received ethical approval from the North Node Privacy Advisory Committee as detailed above. This research was conducted in a safe haven which adheres to the principles and standards outlined in the Charter of Safe Havens in Scotland [40] to ensure patient identity and privacy was protected. All research methods using human data were performed in accordance with the Declaration of Helsinki.

## Acknowledgements

We thank the Health Foundation for providing financial support for this work and for facilitating the Networked Data Lab partnerships which have informed this work. The Health Foundation is an independent charity committed to bringing about better health and healthcare for people in the UK. We thank NHS Grampian and Aberdeen City Council for allowing the use of this data. We thank Hugh Paterson from the council for his contribution in facilitating the sharing of the Aberdeen City Council Child Protection Register data. We also thank the Grampian Data Safe Haven (DaSH) for processing the data used in this study, as well as providing the secure platform used in this analysis and administrative support. We are grateful to members of the public who have made invaluable contributions to this work. This work uses data provided by patients and collected by the NHS as part of their care and support. We thank the anonymous reviewers for their careful reading of this manuscript and for their constructive comments.

## Public Involvement

A detailed summary of the Patient and Public Involvement and Engagement (PPIE) activities conducted in relation to this work to date can be found in a GRIPP-2 reporting tool in the supplementary materials (Table S2). In short, PPIE activities have influenced the design and conduct of this study in a range of ways. Review of lay summaries of the project for data access and ethical approval applications improved readability and comprehension.

Discussions with both the ACHDS PPIE group and the group of professionals have confirmed the importance of the topic, particularly the need for greater integration of a variety of data sources.

Their insights have also shaped the research questions asked and the types of methods applied to answer those questions. Discussions with both groups have informed our description of data management, particularly concerning data protection, in outputs associated with this project. Their insights have also informed the acknowledged limitations of this study.

## Data Availability

All analysis was carried out in the Grampian Data Safe Haven (project ID: DaSH520) on pseudonymised individual-level data. As per the Scottish Safe Haven Charter, only aggregate data can be released from the Grampian Data Safe Haven for publication, but all individual-level data will be archived for 5 years on project completion and may be accessed by application to the Grampian Data Safe Haven (email dash{at}abdn.ac.uk) on condition that appropriate project approvals are secured. Following the archiving period, this data will be deleted.

Data analysis and figure generation were conducted using R (version 4.2.1) in RStudio (version 2023.06.2 build 561). Summary data supporting the findings and visualisations included in this work are available as supplementary materials of this work and in the project GitHub repository (URL: https://github.com/will-ball/NDL3_Child_Protection)

## Abbreviations

CPS: Child Protection Services
CPR: Child Protection Register
CPP: Child Protection Plan
ACCCPR: Aberdeen City Council Child Protection Register
CAMHS: Child and Adolescent Mental Health Services
PIS: Prescribing Information System
CHI: Community Health Index
DaSH: Grampian Data Safe Haven
TRE: Trusted Research Environment
NHS: National Health Service
SIMD: Scottish Index of Multiple Deprivation

