## Supplementary Table 1. CRediT authorship statement for "Serious mental health diagnoses in children on the Child Protection Register: a record linkage study"

**Table S1** CRediT Statement

| Term | Description | Authors |
|---|---|---|
| Conceptualization | https://credit.niso.org/contributor-roles/conceptualization/ | All authors |
| Data curation | https://credit.niso.org/contributor-roles/data-curation/ | HR, ML, WB, JB, KO'S on behalf of Grampian DaSH, CA/MM on behalf of ACC |
| Formal analysis | https://credit.niso.org/contributor-roles/formal-analysis/ | WB, JB |
| Funding acquisition | https://credit.niso.org/contributor-roles/funding-acquisition/ | JB, CB, SG |
| Investigation | https://credit.niso.org/contributor-roles/investigation/ | WB, JB |
| Methodology | https://credit.niso.org/contributor-roles/methodology/ | WB, JB, CB |
| Project administration | https://credit.niso.org/contributor-roles/project-administration/ | JB, CB |
| Resources | https://credit.niso.org/contributor-roles/resources/ | CA, MM, KO'S |
| Software | https://credit.niso.org/contributor-roles/software/ | WB, JB |
| Supervision | https://credit.niso.org/contributor-roles/supervision/ | JB, CB, CA, MM |
| Validation | https://credit.niso.org/contributor-roles/validation/ | JB, WB |
| Visualization | https://credit.niso.org/contributor-roles/visualization/ | WB, JB |
| Writing – original draft | https://credit.niso.org/contributor-roles/writing-original-draft/ | WB |
| Writing – review & editing | https://credit.niso.org/contributor-roles/writing-review-editing/ | All authors |
| Patient and Public Engagement | | MRD, SG, BO, WB, JB |

| Author | Initials |
|---|---|
| William P Ball | WB |
| Caroline Anderson | CA |
| Corri Black | CB |
| Sharon Gordon | SG |
| Michael Lackenby | ML |
| Martin Murchie | MM |
| Bārbala Ostrovska | BO |
| Katherine O’Sullivan | KO'S |
| Helen Rowlands | HR |
| Magdalena Rzewuska Díaz | MRD |
| Jessica E Butler | JB |
