## Supplementary Table 2. GRIPP2 PPIE Reporting for "Serious mental health diagnoses in children on the Child Protection Register: a record linkage study"

**Mental health prescribing and specialist outpatient referrals for children at risk of harm: An analysis of the Aberdeen City Council Child Protection Register and electronic health records.**

**Table S2**

Patient and public involvement (PPI) in this study, described using the GRIPP 2-SF checklist (Staniszewska et al., 2017)

| Section and Topic | Item |
|---|---|
| 1. **Aim**<br>Report the aim of the study | We wanted to look at the mental health care service use of children listed on the Aberdeen City Council Child Protection Register (between 2012 and 2022) using healthcare administrative information on community prescriptions and referrals to outpatient Child and Adolescent Mental Health Services (CAMHS) for the wider NHS Grampian region.<br><br>We followed a process which let us connect these sources of information together for specific individuals. We did this in a way that meant that sensitive information would be accessed safely by researchers who could not see names, dates of birth or addresses alongside their health information.<br><br>We wanted to understand which types of people were listed on the child protection register and count how many mental health outcomes they had compared to people not on the register. As children on the register had differences in their age compared with those not on the register, we also calculated rates to allow for better comparisons between the groups. |
| 2. **Methods**<br>Provide a clear description of the methods used for PPI in the study | Our approach to PPI methods has previously been described in more detail (table S1 from Ball et al., 2022), but in short involved the development of a framework based on the National Institute for Health and Care Research (NIHR) guidance. This included working alongside the funder of this work and the creation of a local PPI plan for specific activities and has led to the establishment of the ACHDS PPIE group. This group includes people from the local community in the North East of Scotland from a variety of backgrounds.<br><br>In addition to activities undertaken with the core members of the ACHDS PPIE group, we also held online discussions with a group of 16 professionals who interact with and support young people who are vulnerable. This included social workers, people working in schools (primary and secondary) and from the third sector (i.e. charities that focus on youth work).<br><br>We intend that these groups will contribute to this research at all stages of project development and execution. To date, these groups have assisted in refining the focus of research questions and developing analysis plans. One member of the ACHDS PPIE group has contributed review and editing for this pre-print article and has been listed as a co-author. Prior to final publication of these results in a peer-reviewed academic journal we will also involve these groups in more thorough sense-checking and interpretation of results, as well as further checking of outputs for readability/comprehension. |
| 3. **Results**<br>Outcomes—Report the results of PPI in the | <u>Following presentation by analytical team of the early research questions and analysis plans, and group discussion the ACHDS PPIE group:</u> |

| Section and Topic | Item |
| --- | --- |
| study, including both positive and negative outcomes | • Highlighted the importance of understanding how children came onto the register and the context of efforts to avoid a registration.<br>• Advised that we should carefully consider the limitations of the available (administrative) data.<br>• Stressed the importance of considering readers of research outputs who might have lived experience of interactions with social work teams or being on the child protection register.<br>• Recommended that we clearly communicate the steps that have been taken to protect data about individuals in order to reassure the public that their information has been handled in a responsible and ethical manner.<br><br><u>Following presentation by the analytical team of their proposed analytical plans, and group discussion, the group of professionals who support children experiencing vulnerability:</u><br><br>• Highlighted the importance of timely and accessible sharing of information about vulnerable children. This took the form of personal experiences and opinions of information sharing/collaboration in practice between professional groups. Various real-world examples were shared of times when information sharing broke down or was less than optimal, leading to negative outcomes for children which could have been avoided.<br>• Suggested additional sources of information that could provide a more rounded contextual overview of children experiencing vulnerability.<br>• Raised that the Child Protection Register listed mostly younger children and that the long-term impacts of their early life experiences on their mental health may not manifest until later.<br>• Discussed the concept of vulnerability, specifically noting that children on the child protection register are not the only ones who experience vulnerability. |
| **4. Discussion**<br>Outcomes—Comment on the extent to which PPI influenced the study overall. Describe positive and negative effects | To date PPI activities have influenced the design and conduct of this study in a range of ways. Review of lay summaries of the project for data access and ethical approval applications has improved readability and comprehension. Discussions with both the ACHDS PPIE group and the group of professionals has confirmed the importance of the topic, particularly the need for greater integration of a variety of data sources. Their insights have also shaped the research questions asked and the types of methods applied to answer those questions.<br><br>Discussions with both groups have informed our description of data management, particularly in relation to data protection, in outputs associated with this project. Their insights have also informed the acknowledged limitations of this study. |
| **5. Reflections**<br>Critical perspective—Comment critically on the study, reflecting on the things that went | Speaking to a range of professional and non-professional participants has provided valuable perspectives on a range of topics related to this work. Speaking to the non-professionals in the ACHDS PPIE group has been rewarding due to the group being more established and embedded in the work of this research team. They have a good familiarity with the researchers and background knowledge provided by interaction related to previous projects on similar topics. |

**Mental health prescribing and specialist outpatient referrals for children at risk of harm: An analysis of the Aberdeen City Council Child Protection Register and electronic health records.**

| Section and Topic | Item |
|---|---|
| well and those that did not, so others can learn from this experience | External pressures on timelines and delays in the project due to data access have resulted in publication of this pre-print prior to completion of all intended PPI activities. We intend to conduct PPI activities to inform our interpretation and framing of results, followed by an updated article which incorporates emerging insights. |
